# Gaps in integration of sexual and reproductive health and rights into climate change research in sub-Saharan Africa: A scoping review

**DOI:** 10.1101/2025.02.26.25322673

**Authors:** Jacinter A. Amadi, George Odwe, Francis O. Onyango, Beth Kangwana

## Abstract

Sub-Saharan Africa is faced with triple challenges of high vulnerability to climate change (CC) impacts, high levels of inequality and poor sexual and reproductive health and rights (SRHR) outcomes. Climate change can worsen SRHR situation for high-risk groups such as women, children, adolescent girls and people living with HIV. This scoping review takes stock of the state of research on the intersection between CC and SRHR in SSA with a view of identifying gaps and opportunities for effective evidence generation and integration in climate actions. The review followed Arksey and O’Malley framework. Data charting was conducted using Population, Exposure, Comparator, Outcome tool in Covidence. Thirty-seven (37) studies were reviewed, 57% were quantitative while 41% and 2% used qualitative and mixed methods respectively. SRHR components investigated include maternal newborn and child health at 43% (n=16), HIV at 19% (n=7), gender-based violence at 11% (n=4), and fertility intentions and outcomes at 11% (n=4). SRHR elements that are under-researched in the context of CC include access to and uptake of contraceptives, post abortion care, harmful practices (child marriages and female genital mutilation), menstrual health, pregnancy losses, bodily autonomy, and (in)fertility. Drought, floods, heat stress and rainfall seasonality have received fair attention in research, yet impacts of climate hazards like tropical cyclones, wildfires and salt-water intrusion are missing in research. There is inadequate research at the intersection of CC and SRHR hindering progress towards climate resilience and attainment of good health and well-being for all. Effective and equitable integration requires that SRHR issues be recognized, and deliberate investments (research, policies, programs, interventions and financing) put in place to address the critical SRHR gaps and climate vulnerabilities to enhance resilience.

## Introduction

Impacts of climate change (CC) on human systems continue to increase in unprecedented ways and magnitude [1]. Climate-related events such as severe droughts, heavy rainfall, floods, cyclones, heatwaves, wildfires, and saltwater intrusion cause morbidity and mortality affecting millions of people globally [1]. Sub-Saharan Africa (SSA) region is worst affected by the vagaries of CC. The region is characterized by acute water stress, food insecurity, and climate-induced diseases, especially in areas with high population density and poverty rates [2]. Climate change impacts such as heat stress, rainfall variability and drought are common in the West African region, and have been linked to persistent food insecurity and water-related challenges [2]. Similarly, the East and Southern Africa region experiences extreme weather events (EWE) such as drought and floods, cyclones and rising temperatures, which have been associated with adverse impacts on water availability, livelihoods, food security and health [2]. Other notable impacts of CC in SSA include loss of habitation, forced displacements and unplanned migrations, infrastructural damages, inaccessible health facilities, disrupted health services, and deteriorating mental health [1,3]. It is noteworthy that these CC impacts disproportionately affect countries or regions that contribute the least to climate change [3,4]. Additionally, vulnerabilities to climate-related risks vary by gender, age, religion, socioeconomic, and disability status [5].

Climate change has the potential to reverse gains made in the health sector. It can exacerbate unmet need for sexual and reproductive health and rights (SRHR), especially in low- and middle-income countries due to higher levels of poverty and income inequality, and weak healthcare systems [6]. Sexual and reproductive health is a state of physical, emotional, mental, and social wellbeing in relation to all aspects of sexuality and reproduction, not merely the absence of disease, dysfunction, or infirmity [7]. Core elements of SRHR include maternal, newborn and child health (MNCH); safe abortion services; family planning; prevention and management of sexually transmitted infections (STIs) including human immunodeficiency virus (HIV); prevention and management of infertility; prevention and management of cancers of the reproductive system; detecting and preventing Gender Based Violence (GBV), forced marriage, transactional sex, and sex trafficking. Sub-Saharan Africa has made great progress in SRHR over the past two decades. The gains include a 33% reduction in maternal mortality, increased coverage of child immunizations, decline in malaria-related child mortality, a drop in HIV incidence, and a two-fold increase in contraceptive uptake [8]. Therefore, if not taken into consideration, climate change can water down the gains made in SRHR.

Impacts of CC disproportionately affect the most vulnerable populations. In SSA, women, children and adolescents, particularly girls, face heightened vulnerability due to the impacts of climate risks [1]. Literature shows that climate extremes directly affect women and girls’ health by increasing the risk of maternal or infant injury or death, disrupting access to family planning and HIV care, decreasing reproductive autonomy and resulting in poor menstrual hygiene [9]. Climate-related extreme events may also have an indirect impact on maternal and newborn health, for example, adverse birth outcomes such as preterm births and low birth weights, increased GBV including intimate partner violence (IPV), increased HIV transmission rates [9], increased unintended pregnancies, and unsafe abortions [10]. Given Africa’s high vulnerability to climate change, its low adaptive capacity, and existing high levels of inequality, the already critical state of SRHR in the region will likely deteriorate further [1]. Addressing this challenge requires efforts that focus on addressing the critical SRHR-CC intersection gaps and empowering women and girls to make informed decisions and choices about their sexual and reproductive health.

Achieving universal health care, including SRHR is a major pathway for individual, community, and institutional climate resilience [11]. This can occur through increased access to and uptake of modern contraceptives, better antenatal care services, improved post abortion care, and reduced SGBV and HIV transmission. These factors improve women’s and girls’ health and enhance their control and decision-making powers over sexual and reproductive health matters [12]. Consequently, improved access to SRHR increases women’s and girls’ ability to engage in empowerment activities like education, income generation, employment opportunities, and access to resources [9], factors that promote climate resilience. Further, access to child health services like vaccinations and preventive care can reduce vulnerabilities of at-risk populations to climate-related diseases like malaria and diarrhea that afflict millions of children in the region. Therefore, identifying and reducing unmet need for SRHR contributes to climate change resilience and improves communities’ and health systems’ adaptive capacity through improved health and well-being of women, girls and children, including increased access to health services, education, nutrition, and family planning [13].

It was until 2021 that health was brought at the center of global climate discussions during United Nations Framework Convention on Climate Change (UNFCCC) Conference of Parties (COP26). Over the years, CC change research and funding have been skewed towards sectors such as agriculture, water, and energy with health sector getting minimal attention. Furthermore, within health research, focus has been on climate-sensitive and infectious diseases such as malaria and cholera [14,15], other health aspects such as SRHR remaining under documented. In 2023, United Nations called for accelerated research on MNCH [16], one of the key elements of SRHR. The recent occurrences in global climate change spaces point to the gaps in CC and SRHR intersection research that need be urgently addressed. Therefore, this scoping review takes stock of the state of research on the intersection between CC impacts, and SRHR outcomes in sub-Sahara Africa and identifies gaps and opportunities for effective SRHR-CC evidence generation, and integration in climate change and health policies, plans, and interventions at all levels. We analyze published documents on the impacts of climate change on SRHR to explore the state of research at the intersection and explore gaps and opportunities for their integration in the region.

## Materials and methods

### Database search

The study identified publications integrating SRHR in climate change research in SSA using Arksey and O’Malley framework for scoping reviews [17]. Electronic databases for peer reviewed journal articles and reports were searched to identify relevant literature using MyLOFT with the support of Kenyatta University Librarians. A first search of journal databases including Pubmed, Elsevier, Springer, Wiley, Taylor and Francis, Oxford Academic Journals, African Journals Online, ScienceDirect, JSTOR, and Saje Journals was done in May 2024 using CC and SRHR key words (S1 Table). The search was restricted to the period between January 2010 to April 2024 consistent with availability of evidence. A second search was conducted in PubMed (S2 Table) using specific SRHR terms derived from Starrs [7] such as maternal, newborn, and child health; abortion care; family planning; HIV; infertility; cancers of the reproductive system; gender based violence including intimate partner violence; forced or early marriage, transactional sex and sex trafficking in addition to climate change search terms to identify additional studies investigating impacts of climate change on various SRHR components that might have been left out during first search. Peer-reviewed journal articles at the intersection of climate change and individual components of SRHR were included. The accessed documents were analyzed to identify the state of SRHR-CC integration in research and gaps and potential entry points for comprehensive integration.

### Search strategy and selection criteria

The search strategy used key words and subject headings relating to climate change and SRHR based on definition provided in this review. Reports generated from the search were managed in Mendeley online library (https://www.mendeley.com/search/) and then uploaded in Covidence (https://www.covidence.org/) for analysis. Records were subjected to first stage screening by a single reviewer using laid-out inclusion and exclusion criteria. Selection and screening of journal articles and reports followed the checklist recommended by Preferred Reporting Items for Systematic Review and Meta-Analysis extension for scoping reviews (PRISMA-ScR) [18]. Records having qualitative, quantitative, mixed method approaches, short communications, commentaries, reviews, and meta-analyses were analyzed in the first stage of the review. Studies were included in the full-text review if (i) they were on climate change (including climate extremes, events, and disaster) and mentioned at least one aspect of SRHR; (ii) they were on SRHR or its components and mentioned climate change hazards (iii) if they were conducted in SSA region and those published in English. Documents were excluded if they solely explored climate change or SRHR and if they were not conducted in the region. Reviews, commentaries, abstracts, reproductive studies on animals and plants and publications without full-text articles were also excluded. Excluded reports did not meet the data charting criteria.

### Data charting and synthesis

Data charting was conducted using a data extraction template developed following the Population, Exposure, Comparator, Outcome (PECO) framework in Covidence. Data extraction template was pretested, and adjustments made before commencing the data extraction process. Data extracted included study details (author, year of publication, setting), data source, data collection method, climate change event(s), study population, impacts of climate event(s), SRHR element(s), and SRHR outcomes. Data extraction, review and synthesis were conducted sequentially by a single reviewer. Data synthesis followed a thematic analysis of SRHR domains. PRISMA-2020 flowchart was used to illustrate article selection process.

The search yielded 8822 records. A total of 7589 ineligible and 735 duplicate reports were removed. Four hundred ninety-eight (498) articles were screened by title and abstract followed by full-text assessment of 117 articles for eligibility. Thirty-seven (37) articles were retained for data extraction (Fig 1). Some excluded articles were reviews and meta-analysis [19–21], commentary or editorial [22–24], while others did not include both CC and SRHR in their content [25–28].

**Fig 1.**
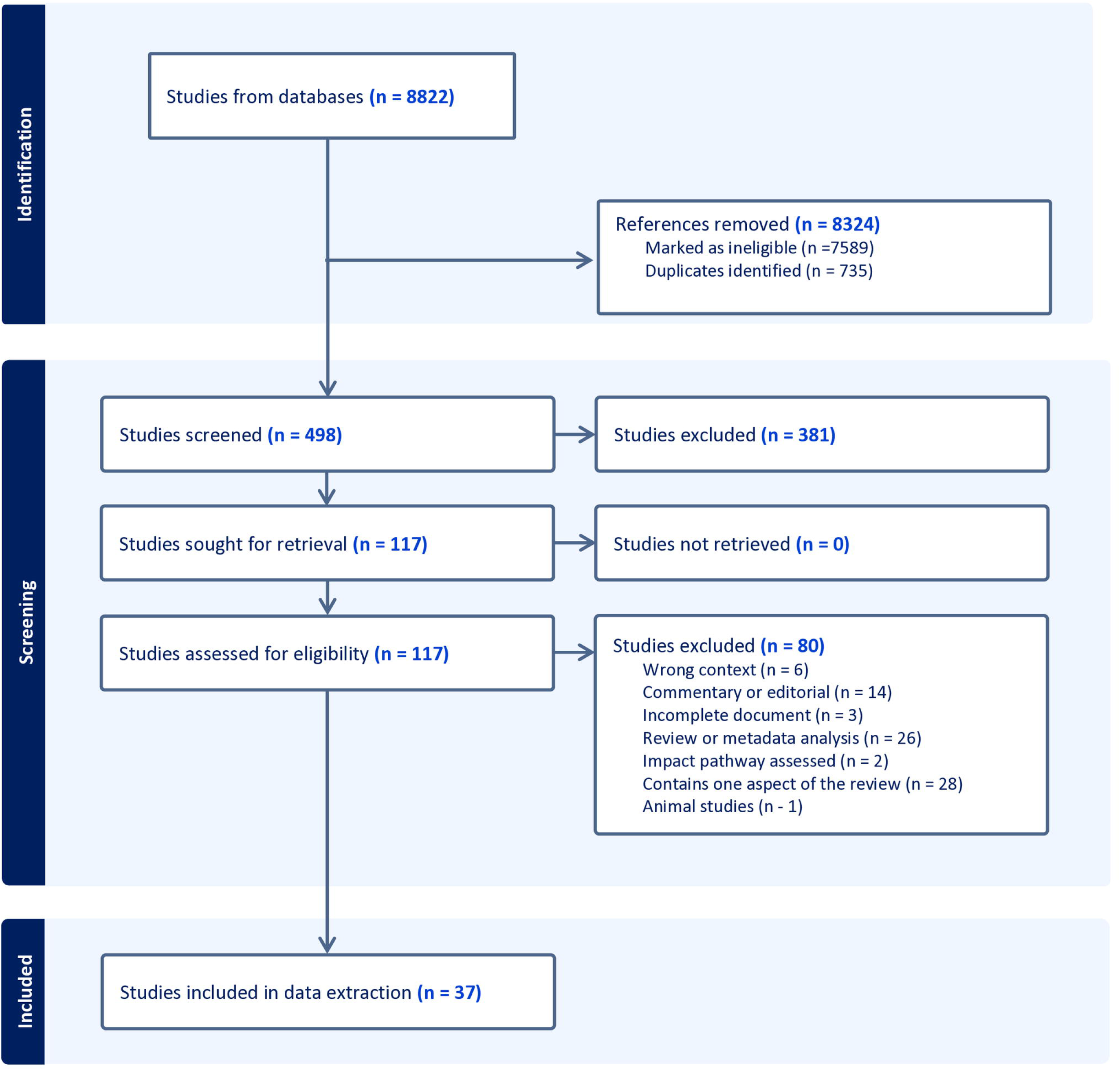
PRISMA 2020 flow chart showing document screening and eligibility assessment process.

### Ethical consideration

The study did not require ethical approval as it did not entail research on human subjects.

## Results

### Study characteristics

S3 Table summarized characteristics of 37 studies included in the review. Majority of the studies (81%, n=30) were published after the year 2020. Fifty seven percent (57%, n= 21) were quantitative studies while 41% (n=15) were qualitative studies and one study used mixed methods approach. Quantitative studies applied cross-sectional, case control, case series, longitudinal, and cohort study designs. Table 1 shows the distribution of studies at the intersection between CC and SRHR by country in the SSA region. Four articles were regional studies [29–32] while three were global studies with reference to SSA [33–35]. The SRHR components investigated were MNCH (43%; n=16), HIV (19%, n=7), GBV including IPV and violence against women and girls (VAWG) (11%; n=4), and fertility intentions and outcomes (11%; n=4). Sixteen percent (16%; n=6) of the articles were studies on multiple SRHR elements including MNCH combined with fertility and HIV [36]; HIV and GBV [37]; early marriage, MNCH and FP[38]; HIV, GBV and MNCH [39], and MNCH and GBV [40]. Impacts of extreme heat, drought, changing rainfall patterns and floods were the main climate hazards studied in 24%, 24%, 14% and 11% of the articles respectively. Other studies either combined multiple CC risks (19%) or assessed climate change impact collectively (8%).

**Table 1.**
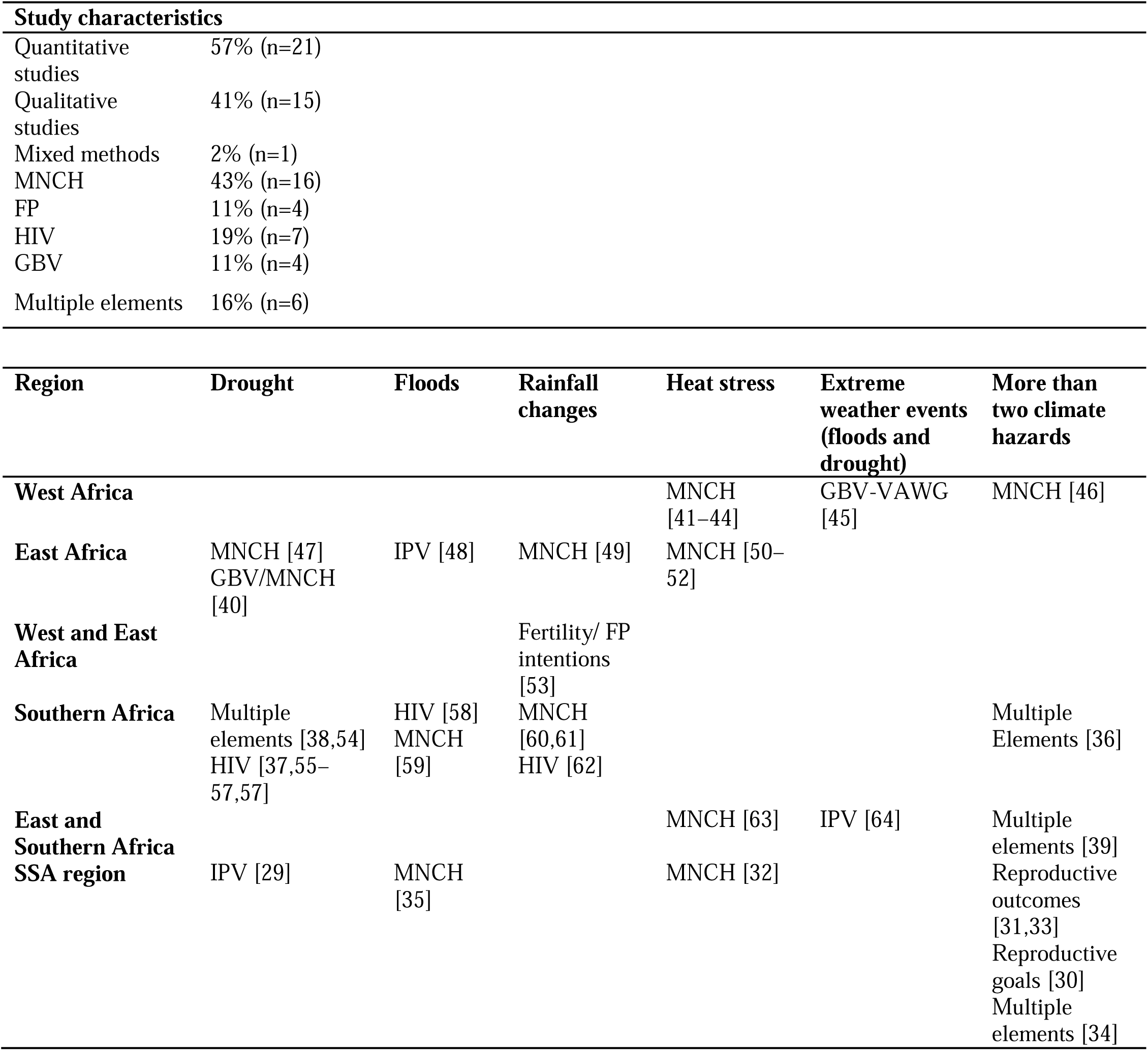
Characteristics of studies included in the review.

### Maternal newborn and child health

The metrices assessed under MNCH included child health outcomes [46,47,63], antenatal health and experiences [35,49,50,60], knowledge levels [41], access to healthcare [59,61], breastfeeding and childcare practices [42], and interventions to address adverse effects of climate-related events [51] (Fig 2). Studies showed that exposure to heat stress during the third trimester of pregnancy reduces birthweight [46,63]. The length and strength of heat waves adversely affects birthweight. Ambient temperatures greater than 35°C were associated with increased odds of wasting and underweight among children under 5 years [32]. A positive impact of higher seasonal Normalized Difference Vegetation Index (NDVI) on birthweight was reported in Mali [46]. In Kenya, a study investigating early gestational exposure to severe drought on child health outcomes showed that exposed children had lower body weight compared to non-exposed siblings [47]. Food insecurity was a strong CC impact pathway affecting maternal health and child health outcomes. In Uganda, indigenous women had the greatest negative maternal-infant health outcomes resulting from CC-related food insecurity compared to non-indigenous women [49].

**Fig 2.**
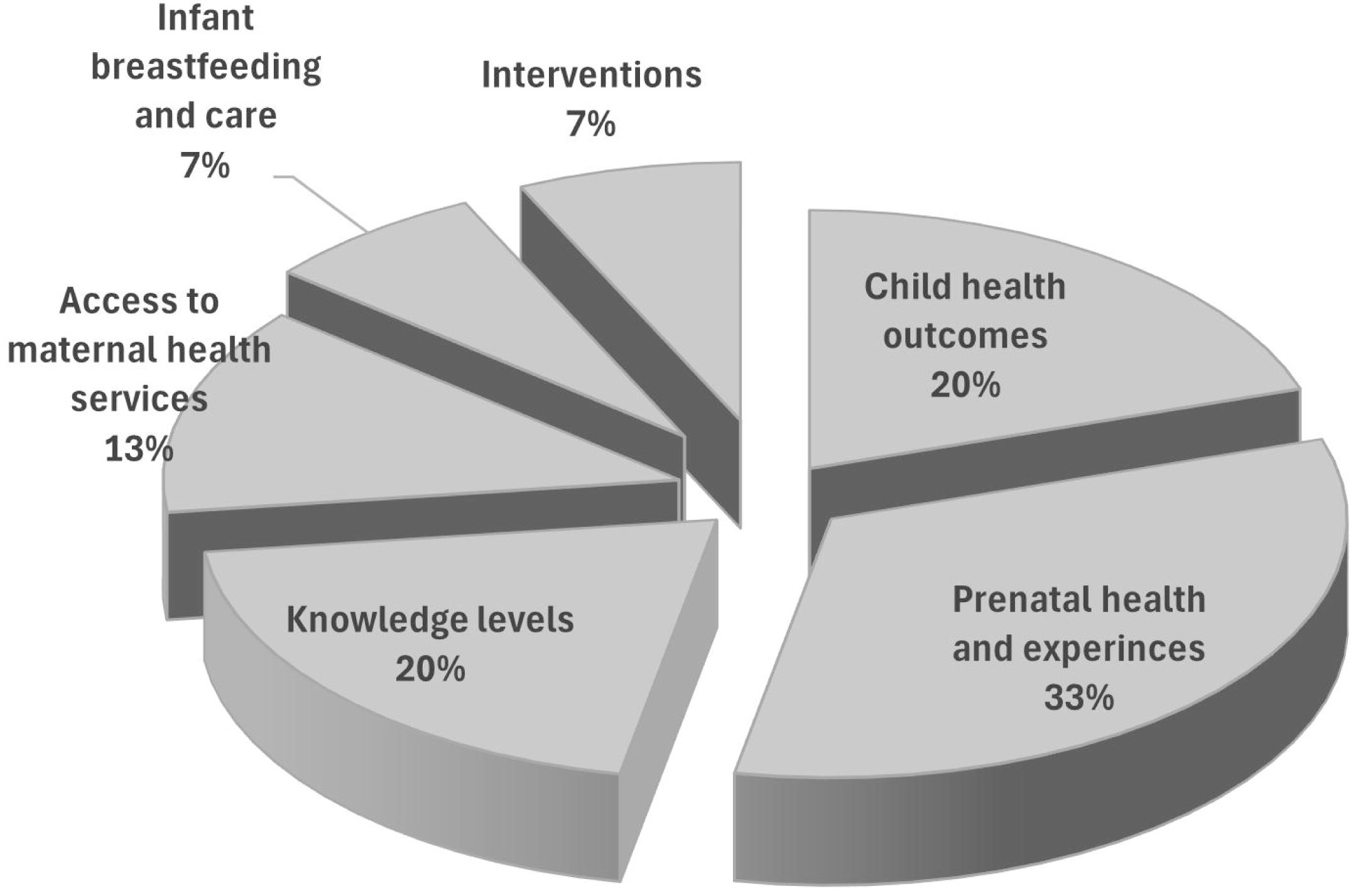
Attributes investigated under Maternal Newborn and Child Health (MNCH) component.

Studies in Zambia and Mozambique investigated the influence of floods [59] and rainfall seasonality [41] on access to and utilization of maternal health services. Floods were found to hinder access to maternal health services. Similarly, low numbers of institutional deliveries were reported to occur during rainy season. The effect was greater for deliveries compared with antenatal care (ANC) visits since women were isolated from accessing delivery facilities. A study conducted in South Africa showed that rainfall seasonality affected ANC attendance with lowest visits ≤ 4 occurring during rainy seasons [60]. The rains not only cause infrastructural damage and limit access to health facilities but also correspond to low food availability and increased agricultural labor, factors that are associated with adverse birth outcomes such as reduced birth weight and head circumference [60]. Another study analyzing impact of floods found that exposure to floods during pregnancy increased the risk of pregnancy loss due to injuries or trauma following flood events. The risk was greater for women outside peak reproductive age (<21 years) and among those in rural areas [35]. A study conducted in Kenya documented the lived experiences of pregnant women under extreme heat [50]. The study showed that heat stress disrupts social and interpersonal relations reducing quality of selfcare and childcare.

Studies exploring perceptions and knowledge about health impacts of extreme heat on MNCH reported lack of awareness among women in Burkina Faso [43] and communities in Kenya [51]. In Kenya, high ambient temperature was associated with early introduction of alternative foods to infants younger than 6 months and reduction in Kangaroo care [52]. Similarly, extreme heat was reported as a barrier to exclusive breastfeeding and a cause of early introduction of supplemental fluids for infants under 6 months [42].

### Gender-based violence

Studies investigated the influence of floods and drought on intimate partner violence (IPV) and VAWG. A study in rural Kenya found that women in agriculture had greatest risk of experiencing physical and sexual violence compared to urban counterparts when exposed to floods [48]. A similar study exploring the relationship between extreme weather and IPV in Uganda, Zimbabwe and Mozambique reported that EWE such as floods and drought increase violence against women and girls by affecting their access to income and employment [64]. Economic burden and displacements that result from EWE are the underlying factors that cause high prevalence of IPV. Another study found that severe drought was associated with greater risk of sexual violence [29]. The risk of violence increased with severity of drought and frequency of floods, an occurrence attributed to increased conflicts in households experiencing soring commodity prices and scarcity of resources. A study in Chad examined the association between violence and women’s resilience to climatic events and found that violence exposes women and girls to adverse consequences such as reproductive health injuries and morbidities [45]. The study further reported that socio-cultural practices hinder women from fully participating in decision-making processes and exercising control over financial resources, thereby reducing their capacity to cope with crises. Climate change impacts limit access to care and reduces resources needed for survival, thus widening SRH gaps for women and girls.

### Sexually transmitted infections including HIV

Research on HIV has been going on for decades, however, studies integrating impacts of CC risks are more recent as revealed in this review. Studies reviewed focused on HIV prevention, management and service provision, with drought being the main CC risk investigated followed by floods and changes in rainfall trends. HIV dimensions examined were transactional sex and HIV prevalence [56], HIV prevention, management and care [37,55], adverse HIV outcomes on people on anti-retroviral therapy (ART) [62], institutional capacities to provide HIV services and care [57], climate impact on PLHIV and service provision [58,65]. A study in Malawi reported that drought increases HIV prevalence by 15%, with the effect being mediated by risky sexual behavior like transactional sex [56]. Transactional sex is a key exposure pathway between climate change and HIV infections [37,56]. A similar study in Lesotho found that living in drought stricken areas was associated with higher HIV incidence among young female (aged 15-24) [37]. A report from South Africa found that drought negatively affected HIV treatment adherence by disrupting income, livelihoods and food systems, thus, increasing the risk of ill-health [55,65] and higher odds of unsuppressed viral load. The adverse HIV outcomes were accompanied by high mortality among PLHIV on ART and exacerbated by depressed rainfall or drought [62]. A study in Namibia reported that floods negatively impacted HIV service provision and care by limiting access to facilities, leading to reduced HIV testing, thereby weakening prevention of mother to child transmissions (PMTCT) efforts, and disrupting access to treatment [58].

### Family planning

Studies analyzing the impacts of climate change on reproductive health (RH) were regional [30,31] and national in scope [53,54]. This review did not find studies that directly assessed the impacts of CC on access to and uptake of family planning (FP) services in the SSA region. However, fertility intentions and reproductive goals [30,53], life course transition [54], and inclusion of RH in policies [31,33] were the FP proxies investigated. Studies reported an association between a good growing season and increased desire to bear children in future and a higher likelihood of FP discontinuation [53]. In contrast, exposure to extreme temperature was associated with low fertility preference and low ideal family size [30]. Studies showed that fertility goals varied across SSA region and among populations in response to CC hazards. In Malawi, exposure to drought increased adolescent’s transition into first birth and young women’s transition into cohabitations and marriages, behaviors linked to resource constraint (food and income) experienced during drought [54]. A study assessing the status of inclusion of RH in climate change policies in Africa reported that RH was not adequately recognized or included in climate change adaptation strategies [31]. Another study reported that only 14% of countries in SSA included actions and strategies to slow population growth in the Nationally Determined Contributions (NDCs) submitted in 2020 and that data scarcity is an impediment to FP reporting and integration into policy [33].

### Early marriages, risky sexual behaviors and other SRHR components

Only two (2) studies analyzed the impact of climate change on early marriages. In Zambia, economic consequences of drought such as reduced household income and food insecurity increased women’s vulnerability to transactional sex and early child marriages [38]. In Malawi, a study reported early sexual debut, and accelerated transition into unions among adolescents and young women exposed to drought [54]. Crop yield reductions, psychosocial stress, limited food and livelihood resources were the factors that accelerated early marriage and childbearing among young women (under 24 years). A study assessing health consequences of large-scale displacements observed that CC is not adequately addressed in health care planning and proposed the need for robust political and governance solutions to health needs in humanitarian setting [40]. This review found a dearth of information on the impacts of CC on harmful practices like female genital mutilation (FGM) and other SRHR elements like menstrual health, sex trafficking, infertility, abortion care, and cancers of the reproductive system.

## Discussion

This scoping review provides a synthesis of the impacts of climate change on SRHR outcomes in SSA region. The review documents studies conducted on the intersection, identifies gaps and proposes recommendations for effective integration of SRHR in CC research policy and interventions for climate resilience. In relation to climate change research, maternal, newborn and child health (MNCH) was the most researched SRHR component (n=16) followed by HIV prevention and care (n=7), reproductive goals (n=4), and GBV (n=4). A recent study conducted in the LMICs corroborates this finding [66]. The focus on MNCH and HIV may be related to the high-level political commitments to reducing maternal and under-five mortality as well as the global strategy on HIV prevention and response. Drought, heat stress, changing rainfall seasons and floods were the most studied climate change events. Studies reviewed investigated direct impacts and exposure pathways of climate change on SRHR outcomes. Direct impacts included effects on access to maternal and child health services, effects on biological mechanisms modulating SRHR outcomes, heat stress effects, and immediate and long-term physical and mental health impacts. Exposure pathways assessed were food insecurity, diminishing livelihood resources, undernutrition, increasing commodity (e.g. food) prices, economic shocks, displacements and migrations. Adverse SRHR outcomes resulting from climate risks examined included changing fertility goals and choices, poor prenatal and child health, disrupted service delivery, changing sexual behavior patterns, adverse birth outcomes like still births, pre-term births and low birth weight, increasing physical and sexual violence, early sexual debut and early entry into unions, human morbidity, and mortality.

This scoping review showed that SRHR has not been effectively and equitably addressed in climate change research in SSA. There is a narrow range of SRHR components in the studies reviewed; much of SRHR research is still siloed while CC policy and actions hardly recognize or largely exclude SRHR issues. Neonatal and child health outcomes, antenatal health and experiences, awareness and knowledge levels, access to maternal health services, and neonatal-care and self-care were the dimensions investigated under MNCH. Biological mechanisms by which drought and heat stress impact maternal and child health included changes in gene function [47] and reduction in placental blood flow [44] causing adverse birth outcomes like reduced gestational age and increased chances of pre-term birth [63]. Behavioral outcomes like reduced ANC visits and facility-based deliveries [38] were additional ways that led to adverse birth outcomes. Although drought ranks third among climate disaster risks in Africa [67], it causes the greatest mortality, affects millions of people, destroys livelihoods, increases food insecurity, and reduces income [68] factors that negatively affect maternal and child health outcomes. Heat stress poses a potential threat to health and wellbeing of pregnant women and infants and can weaken gains made in reducing maternal and neonatal mortality and morbidity in the region. Normalized Difference Vegetation Index, a substitute for food security status positively corelates with birth outcomes. Thus, seasonal NDVI can be incorporated in early warning systems to trigger early actions and interventions such as supplementation for pregnant women and children [69].

The review further showed that sexual and physical violence continue to be perpetrated on young women in diverse settings, with rural women bearing a heavy burden compared to their urban counterparts [70]. Most studies reviewed focused on IPV, with violence meted by non-partners and violence against adolescent girls remaining understudied. Economic loses and forced displacement are the pathways through which EWE perpetuate violence against women and girls. Rural women, especially those in agriculture, were at the greatest risk of experiencing physical and sexual violence [71]. Socio-cultural practices [72], political, and economic barriers [73] aggravate GBV and gender inequality, and should thus be addressed together in climate actions.

Individuals facing EWE have higher likelihood of exposure to HIV infection [22]. On the one hand, drought increases HIV incidence by contributing to risky sexual behaviors and affecting HIV treatment adherence. On the other hand, floods disrupt HIV service provision and care by limiting access to facilities, HIV testing and treatment [74]. Consequently, understanding the relationship between EWE and transmission of HIV can better position HIV prevention services and programs such as pre-exposure prophylaxis (PrEP) to cater for the needs of at-risk persons who experience these events [22]. Despite the steady progress made on HIV control over the past decades, climate change poses a new threat to the efforts on reducing HIV epidemic globally [75]. Collective actions addressing to both HIV prevention and climate actions will require anticipatory planning while taking into consideration equity, human rights and justice to deliver climate-proof HIV prevention programs, services, care and support in geographic areas under climate crisis [75].

Currently, there are limited country-based studies in SSA evaluating the link between climate change and FP use. While most studies investigated FP aspects such as fertility intentions and reproductive goals, there were no studies linking unmet need for FP and climate risks. Yet, there is great overlap between high fertility, high population growth, high unmet need for FP and high climate vulnerability and risks in many parts of the region [76]. Evidence suggests that CC may worsen factors (socioeconomic, education, mobility, employment, and land tenure) associated with uptake of and access to FP [77]. Recognition of impacts of population growth and inclusion of reproductive health (including FP) in national climate change policies and plans provide a basis for addressing unmet need for FP. Climate change adaptation and health interventions targeted to poor women without formal education can increase the uptake of modern contraceptives and improve SRHR outcomes necessary for climate resilience [78]. Climate change mitigation and adaptation actions should be cognizant of the need to integrate population, health and development needs in the region.

Drought and dwindling rainfall have been associated with increased vulnerability of girls to forced or early marriages and accelerated transition into marriages and childbearing by young women. Poverty, religion, culture, and lack of access to education were the main drivers to child marriage in SSA. Most child marriages occurred in countries across the Sahel, a region ravaged by extreme drought and poverty [79]. Environmental and climate crises are known to worsen drivers of early marriages. It is therefore imperative that actions that tackle child marriages should take into consideration their connection to climate change and vice versa [10].

### Gaps in climate change and SRHR research

Impacts of CC on SRHR outcomes have not been adequately investigated to inform policy and actions. Compared to South Asia and other parts of the world, there are no studies in SSA examining the linkages between saltwater intrusion and maternal and child health outcomes [80–82]. There is also inadequate evidence on impacts of climate hazards on SRH outcomes such as GBV, HIV outcomes (biological and behavioral), access to and uptake of family planning, fertility/infertility, and harmful practices like child marriage and FGM. Climate change risks and subsequent SRHR outcomes are diverse and vary from West to East and Southern Africa thus require contextualization of CC actions and SRHR interventions [30]. The potential impacts of extreme temperatures on male fertility, health of PLHIV, ART use and health-seeking behaviors are some of the understudied SRHR elements that future research should investigate. Furthermore, impacts of CC or EWE on menstrual health, pregnancy losses, bodily autonomy, male infertility, harmful practices (sex trafficking), SRHR outcomes for adolescent girls and minority groups such as indigenous women, persons with disability and sexual minorities are additional dimensions that should be considered in future research. Research should focus on data disaggregation and underserved, marginalized or minority groups. Further still, there is a shortage of data on other climate change hazards such as cyclones, fire weather and air pollution on SRHR outcomes. Mediating factors such as biological mechanisms, behavior changes, and social and economic factors should also be addressed to reveal nuanced relationships with SRHR elements.

Lived experiences of vulnerable persons in high-risk areas should be documented to support policy and actions to address their SRHR needs. Contextualization of SRHR outcomes is needed for informed decision-making and development of effective and appropriate interventions. Research should also explore the impacts of meteorological conditions such as temperature, humidity and solar radiation to identify synergies and combined impacts on SRHR outcomes. A knowledge gap exists on intersecting issues such as culture that exacerbate impacts of CC and widen the gap in addressing SRHR needs. Evidence on factors such as gender, age, and other social, biological and economic characteristics that modulate climate-SRHR impacts are needed to inform the design of targeted interventions for specific groups. Low awareness and knowledge levels among at-risk population like pregnant women, health care providers and communities on health impacts of climate hazards need to be addressed [42,52]. Awareness levels and knowledge on the effects of climate extremes on pregnancy loss [83], and mental health [84] need further investigations. Increased awareness will dispel misinformation on impacts of CC on maternal and child health outcomes. In addition, studies on climate change, migration and SRHR nexus in fragile settings should incorporate displaced or migrant persons’ and host communities’ experiences and engage them in the design of policy and programs for sustainability.

Still, research on CC-SRHR intersection is yet to benefit from the support of global climate funding sources such as adaptation fund, green climate fund, and global environment facility. The funding can be achieved if governments recognize and include SRHR in their national and sub-national policies, plans and programs. Inclusion of SRHR in national adaptation plans, NDCs and other long-term development plans promotes SRHR, wellbeing of all and supports achievement of Sustainable Development Goals SDG 3 and 13. Financing of CC-SRHR interventions should benefit from support from both local and international sources.

### Limitations

This scoping review identified research studies at the intersection between CC and SRHR in sub-Saharan Africa. The search focused on peer-reviewed journal articles published in electronic databases. Due to budget constraints, data extraction was conducted by a single reviewer and the search did not include grey literature, consequently, some documents may have not been captured. The search was also limited to studies published in English leaving out potential studies published in other languages. Given that the topic is nascent, a systematic review may have resulted in exclusion of publications considered not rigorous yet had important insights, thus a scoping review was conducted. Quality assessment was not conducted. This may have limited reporting on certainty of evidences included. Some of the studies reviewed did not incorporate climate data to validate CC in the study locations. In other cases, at risk population such migrants or host communities were not represented among study respondents. The scales at which some of the studies were conducted weakened the application of recommendations due to lack of contextualization and heterogeneity of research settings.

## Conclusions

This scoping review reveals that since 2020 there has been a significant increase in research on the intersection between climate change and SRHR signaling a move towards inclusivity. Majority of the studies are on a narrow range of SRHR components, domains encompassing rights and choices are largely under researched. Drought, floods, heat stress and rainfall seasonality have received a fair attention in climate change research, however, impacts of other climate hazards such as tropical cyclones, fire weather and salt-water intrusion are missing. Intervention research is inadequate hindering progress towards climate resilience and attainment of universal access to SRHR. Consequently, effective and equitable CC-SRHR research integration will require recognition and inclusion of population growth impacts and SRHR needs in national and sub-national climate change policies, plans and actions in SSA, and in the medium-term and long-term national development plans, and integration of climate change in health policies and plans.

## Supporting information

PRISMA-ScR Checklist

## Data Availability

All data supporting the findings of this study are contained in the manuscript and its supplementary materials.

## Acknowledgements

The research team thank Kenyatta University librarians for their support during database search.

## Supporting information

**S1 Table. Search terms used during document screening.**

**S2 Table. Additional search terms used during a second search conducted in PubMed.**

**S3 Table. Characteristics of 37 studies included in the review.**

