## Supplementary Table 1 for "Gaps in integration of sexual and reproductive health and rights into climate change research in sub-Saharan Africa: A scoping review"

**S1 Table: Search terms used during document screening**

| **Key words** | **Search terms** | **Database** |
| --- | --- | --- |
| Climate change, sexual reproductive health, rights Africa | "climate change" OR "climate" AND "change" OR "climate change" OR "extreme weather" OR "extreme" AND "weather" OR "climate crisis" OR "climate" AND "crisis" AND "sexual health" OR "sexual" AND "health" OR "sexual health "OR "sexual" AND "reproductive health" OR "reproductive" AND "health" OR "reproductive health" AND rights AND "Africa" OR "Africa" AND ("2010/01/01"[PubDate]: "2024/04/30"[PubDate] | PubMed  (n=856) |
|  |  | Elsevier  (n=562) |
|  |  | SpringerLink  (n=299) |
|  |  | Wiley online  (n= 2109) |
|  |  | Taylor and Francis  (n= 2213) |
|  |  | Oxford academic journals  (n=242) |
|  |  | AJOL  (N=115) |
|  |  | ScienceDirect  (n=41) |
|  |  | JSTOR  (n=49) |
|  |  | Sage Journals  (49) |
