## Supplementary Table 2 for "Gaps in integration of sexual and reproductive health and rights into climate change research in sub-Saharan Africa: A scoping review"

S2 Table: Additional search terms used during a second search conducted in PubMed

| Database | Key words | Search terms |
| --- | --- | --- |
| PubMed | Climate change, climate disaster, extreme weather, maternal health, newborn care and Africa  (n=1432) | "climate change" OR "climate" AND "change" OR "climate change" AND "climate disaster" OR "climate" AND "disaster" AND "extreme weather" OR "extreme" AND "weather" AND "maternal health" OR "maternal" AND "health" OR "maternal health" AND "infant, newborn" OR "infant" AND "newborn" OR "newborn infant" OR "newborn" AND care AND "Africa" OR "Africa" AND "2010/01/01"[PubDate]: "2024/04/30"[PubDate] |
|  | Climate change, climate disaster, extreme weather, family planning and Africa (n=316) | "climate change" OR "climate" AND "change" OR "climate change" AND "climate disaster" OR "climate" AND "disaster" AND "extreme weather" OR "extreme" AND "weather" AND "family planning services" OR "family" AND "planning" AND "services" OR "family planning services" OR "family" AND "planning" OR "family planning" AND "contraception" OR "contraception" AND "contraceptive agents" OR "contraceptive devices "OR "contraceptive" AND "devices" OR "contraceptive devices" OR "contraceptive" OR "contraceptive agents" OR "contraceptive" AND "agents" OR "contraceptive agents" AND "Africa" OR "Africa" AND "2010/01/01"[PubDate]: "2024/04/30"[PubDate] |
|  | Climate change, climate disaster, extreme weather, Sexual gender-based violence and Africa  (n=261) | "climate change" OR "climate" AND "change" OR "climate change" AND "extreme weather" OR "extreme" AND "weather" OR "climate disaster" AND "climate" OR "disaster" AND "sexual behavior" OR "sexual" AND "behavior" OR "sexual behaviour" AND "sexual" OR "behavior" OR "sexual" AND "gender-based violence" OR "gender-based" AND "violence" OR "gender-based violence" OR "gender" AND "based" AND "violence" OR "gender based violence" OR "intimate partner violence" AND "intimate" OR "partner" OR "violence" AND " Africa" AND "2010/01/01"[PubDate] : "2024/04/30"[PubDate] |
|  | Climate change, climate disaster, extreme weather, forced and early marriage Africa  (n=78) | "climate change" OR "climate" AND "change" OR "climate change" AND "extreme weather" OR "extreme" AND "weather" OR "extreme weather" OR "climate disaster" AND "climate" OR "disaster" AND "forced" AND "marriage" OR early AND "marriage" OR "marriage" AND "Africa" OR "Africa" AND "2010/01/01"[PubDate]: "2024/04/30"[PubDate] |
|  | Climate change, climate disaster, extreme weather, Transactional sex and sex trafficking  (n=5) | "climate change" OR "climate" AND "change" OR "climate change" AND "extreme weather" OR "extreme" AND "weather" OR " climate crisis" AND "climate" OR "crisis" AND transactional AND "sex" OR "sex" AND "human trafficking" OR "human" AND "trafficking" OR "human trafficking" OR "sex" AND "trafficking" OR "sex trafficking" AND ("Africa" OR "Africa" AND ("2010/01/01"[PubDate] : "2024/04/30"[PubDate] |
|  | Climate change, climate disaster, extreme weather, STD including HIV and Africa  (n=193) | "climate change" OR "climate" AND "change" OR "climate change" AND "disasters" OR "disasters" AND "sexually transmitted diseases" OR "sexually" AND "transmitted" AND "diseases" OR "sexually transmitted diseases" AND "HIV" OR "HIV" AND "Africa" OR "Africa" AND "2010/01/01"[PubDate]: "2024/04/30"[PubDate] |
|  | Climate change, climate disaster, extreme weather, cancers of reproductive health  (0) | "climate change" OR "climate" AND "change" AND " extreme weather" OR " extreme" AND " "weather" OR " climate disasters" AND "climate" OR "disasters" AND "reproductive health cancers" OR "reproductive" AND "health" AND "cancers" AND "Africa" OR "Africa" AND "2010/01/01"[PubDate]: "2024/04/30"[PubDate] |
|  | Climate change, climate disaster, extreme weather, Infertility (2) | "climate change" OR "climate" AND "change" AND " extreme weather" OR " extreme" AND " "weather" OR " climate disasters" AND "climate" OR "disasters" AND "infertility" OR "reproductive" AND "infertility" AND "Africa" OR "Africa" AND "2010/01/01"[PubDate]: "2024/04/30"[PubDate] |
