## Supplementary Table 3 for "Gaps in integration of sexual and reproductive health and rights into climate change research in sub-Saharan Africa: A scoping review"

S3 Table: Characteristics of 37 studies included in the scoping review

| **Ref. No.** | **Study (setting)** | **Study design** | **Climate change hazard(s)** | **Study aim** | **SRHR component**  **and (study population)** | **SRHR outcomes** |
| --- | --- | --- | --- | --- | --- | --- |
| 29 | Epstein et al., 2020  (Sub-Sahara Africa) | Cross-sectional | Drought | To assess the association between drought and IPV towards women. | GBV (IPV)  (partnered and married women 15-49 years) | Drought predisposes women to the risk of physical violence. Women at greater risk of sexual violence during severe drought. |
| 30 | Eissler, et al., 2019  (Sub-Sahara Africa) | Longitudinal | Temperature and precipitation anomalies | To analyze the relationship between climatic variability and women’s reproductive goals. | FP  (women of reproductive age) | Higher temperatures are associated with declines in ideal family size (IFS).  Higher precipitation levels have negative short-term effects and positive longer-term effects on IFS. |
| 31 | Mutunga and Hardee, 2010  (Africa) | Qualitative | Not specified | To assess the status of integration of RH/FP in the NAPA process and priority adaptation actions. | Reproductive health  (not specified) | 86% (26/31) of LDCs recognize rapid population growth as a factor that reduces resilience to CC.  19% (5/26) of NAPAs includes RH as adaptation strategies.  7% (2/26) of NAPAs identifies RH projects as part of country's adaptation strategy. |
| 32 | Tusting et al., 2020  (Africa) | Cross-sectional | Heat stress | To investigate the relationship between growth faltering and environmental temperature. | MNCH  (children under 5 years) | Temperature > 35 °C is associated with increase in odds of wasting, underweight and concurrence of stunting with wasting in children. |
| 33 | Dodson et al., 2022  (Global) | Qualitative using content analysis | Not specified | To assess the extent to which countries incorporate impacts of population growth into their NDCs. | FP  (not specified) | 70% of NDCs submitted in 2020 did not include impacts of population growth. Countries with high annual population growth rate did not include impacts of population growth in their NDCs- majority in SSA. Only 14% of population inclusive NDCs have strategies to slow population growth viz through adaptation measures, and national development priorities. |
| 34 | WHO 2023  (Global) | Qualitative study | Not specified | To assess governments' progress in addressing climate change health risks towards achieving the Paris Agreement. | Not specified  (none) | All SSA countries now include health considerations in the submitted NDCs including co-benefits of mitigation, health adaptation and climate finance. |
| 35 | He et al., 2023  (Global) | Retrospective case control | Floods | To analyze the association between gestational exposure to floods and pregnancy loss. | MNCH  (women) | Exposure to floods during pregnancy increases risk of pregnancy loss. The risk is greater among women under 21 and over 35 years) and during 2^nd^ and 3^rd^ trimesters. The risk is higher for women with lower income and levels of education. SSA accounted for about 30% of the pregnancy losses, rural areas having 59% of the losses. |
| 36 | Pullanikkatil et al., 2013  (Malawi) | Case study | Erratic rains, Drought, floods, | To assess local community perceptions about the linkages between population growth, reproductive health, gender, and CC. | FP, Maternal, Newborn and Child Health (MNCH) and HIV  (a youthful rural population) | Gender barriers increase the impacts of climate change and negatively affects adaptation efforts. |
| 37 | Low et al., 2019  (Lesotho) | Cross-sectional | Drought | To determine the association between severe drought and HIV prevalence, behaviors of and care for PLHIV. | HIV  (men and women) | Drought increases the odds of females (aged 15- 24) to engage in early sexual debut and have higher HIV infections. Drought is associated with greater HIV prevalence among young females. Migrations increases chances of young people getting infected with HIV. |
| 38 | Rosen et al., 2021  (Zambia) | Qualitative, Cross-sectional study | Drought | To explore the impact of droughts on gendered livelihood transitions, women’s wellbeing, and their sexual and reproductive health (SRH) outcomes. | FP, Early marriage, MNCH  (adult men and women) | Drought drives changes in fertility intentions as women desire small family sizes. Drought affects accessibility of SRH services (ANC and facility-based deliveries, and contraceptive availability). Drought increases women's vulnerability to transactional sex and early child marriages. |
| 39 | Trummer et al., 2023  (Uganda, Ethiopia, South Sudan,  Zambia,  Zimbabwe) | Qualitative using Webinar | Floods, Drought, Storms, and Heat stress | To analyze the views of experts on the linkage between climate change, migration, and health. | HIV, GBV, MNCH  (stakeholders) | CC worsens SRH outcomes for women and children in humanitarian settings through poor nutrition, limited access to contraceptives, risk of sexual violence and risk of engaging in transactional sex. |
| 40 | Lindvall et al., 2020  (Somalia, Kenya,  Ethiopia) | Qualitative study using in-depth interviews (IDIs) | Drought | To identify knowledge status and gaps regarding health consequences of large-scale displacements. | GBV, MNCH  (Professionals from different agencies) | CC has not been adequately addressed in health care planning.  Robust political and governance solutions for health challenges of migrations and displacement are needed. |
| 41 | Spencer et al., 2022  (Gambia) | Qualitative using IDIs | Heat stress | To explore perceptions of pregnant women on health impacts of occupational heat stress. | MNCH  (pregnant women) | Heat stress worsens pregnancy symptoms and causes heat illness.  Coping strategies to heat stress reduces productivity and can result in negative health impacts e.g. reduced income and quality of life. |
| 42 | Part et al., 2022  (Burkina Faso) | Prospective Cohort  study | Heat stress | To explore the effects of daily outdoor temperature on infant feeding practices and childcare. | MNCH  (pregnant and postpartum women 15-45 years) | Hot climate is a barrier to exclusive breastfeeding and results in increase in supplemental fluids for infants younger than 6 months. Breastfeeding duration decreases as ambient temperature increases for infants older than 3 months. |
| 43 | Kadio 2024  (Burkina Faso) | Qualitative using IDIs, KIIs and FGDs | Heat stress | To assess the perceptions of women on impact of heat stress on physical and mental health | MNCH  (pregnant and postpartum women, their kin and service providers) | Extreme heat affects breastfeeding and reduces child-care (kangaroo care).  Cultural practices can intensify negative effects of heat stress.  There is limited awareness of heat impacts and risks to MNCH among populace and health care providers. |
| 44 | Bonell et al., 2023  (Gambia) | Cohort Study | Heat stress | To evaluate the use of Umbiflow^TM^ in field setting to assess the impact of heat stress on umbilical artery resistance index (RI) | MNCH  (pregnant women) | Resistance index of umbilical artery can explain the association between heat stress and adverse pregnancy outcomes. |
| 45 | Masson et al., 2019  (Chad) | Mixed method using KIIs, DHS data and NGO records | Floods, drought | To establish the linkages between violence and women’s resilience capacities. | GBV  (women) | VAWG prevents survivors from positively managing challenges thus negatively affects their resilience to environmental challenges.  Violence has negative consequences such as reproductive health injuries, and low literacy levels. |
| 46 | Grace et al., 2021  (Mali) | Longitudinal | Rainfall, NDVI and heat stress | To explore pathways linking climate variability to child health outcomes (birth weight). | MNCH  (rural and urban pregnant women) | Higher seasonal NDVI positively impacts birth weight. Exposure to malaria during third trimester corresponded to low birth weights. Women at 3rd trimester are at greater risk of adverse outcomes of heat stress. |
| 47 | Straight et al., 2022  (Kenya) | Quasi-experimental study | Drought | To assess the impact of early gestational exposure to drought on child health outcomes. | MNCH  (children) | Children exposed to severe drought in utero have lower body weight compared to unexposed siblings. Changes in gene function mediate the biological mechanisms for reduced body weight and adiposity in drought exposed children. |
| 48 | Allen et al., 2021  (Kenya) | Cross-sectional study | Floods | To assess the relationship between IPV and severe weather events (SWEs) among Kenyan women | IPV  (Women aged 15-49 years) | Women whose partners work in agriculture have greater odds of reporting IPV and counties that suffer severe flooding have greater odds of physical and sexual violence. |
| 49 | Bryson et al., 2021  (Uganda) | Qualitative  using Focus Group discussion (FGDs) | Rainfall seasonality | To analyze pathways through which CC influences pregnancy among indigenous and non-indigenous women. | MNCH  (rural women) | Food security strongly influences maternal health during pregnancy. Unpredictable weather and dry conditions challenge food availability with great impacts for pregnant women and infants. Indigenous women bear greater burden of negative SRH outcomes. |
| 50 | Scorgie et al., 2023  (Kenya) | Qualitative using IDIs, KIIs and FGDs | Heat stress | To document lived experiences of pregnancy in extreme heat. | MNCH  (pregnant or post-partum women 16 years and above) | Extreme heat is associated with maternal hypertension and tachycardia.  Heat stress disrupts social and interpersonal relations and reduces the ability of women to perform physically demanding labor critical for their survival. |
| 51 | Lusambili et al., 2023  (Kenya) | Qualitative using workshop | Heat stress | To design and development interventions against impact of heat exposure on maternal and neonatal health | MNCH  (pregnant and post-partum women) | Communities are not aware of the heat impacts on maternal and neonatal health. Interventions should be co-designed by diverse stakeholder and be socially and culturally appropriate. Accessibility to water points, education and social behavior change campaigns are effective interventions with high chances of success. |
| 52 | Lusambili et al., 2024  (Kenya) | Qualitative using IDIs, KIIs, and FGDs | Heat stress | To assess community perspectives on the effects of high ambient temperature on the health and well-being of neonates and impacts on post-partum women and infant care. | MNCH  (women, health care providers, and influential community members) | Communities associate high ambient temperatures with insufficient breast milk production and early introduction of alternative foods to infants under 6 months.  Heat stress is also associated with reduced Kangaroo care thus reducing the health benefits to the newborn. |
| 53 | Brooks et al., 2023  (Burkina Faso  Kenya, Uganda) | Cross-sectional study | Rainfall seasonality | To analyze the effect of variations in seasonal agricultural quality on childbearing goals and family planning use. | FP  (women of reproductive age) | Quality of (good) growing season influences the desire for large family size and likelihood of FP discontinuation. |
| 54 | Andriano & Behrman, 2020  (Malawi) | Cross-sectional | Drought | To analyze drought impact on timing, sequencing and characteristics of young women's life course transitions into unions and childbearing. | Fertility intentions  (rural women) | Exposure to growing season’s drought significantly increases young women's transition into first unions (cohabitation and marriages).  Exposure to drought positively influences and accelerates transition into first births among adolescent girls. |
| 55 | Iwuji et al., 2023  (South Africa) | Interrupted time series analysis | Drought | To investigate the relationship between drought and adherence to ART and retention in HIV care. | HIV  (HIV positive individuals 15- 59 years) | Drought significantly decreases ART adherence. Adherence is worse among women aged 15-24 years.  During drought, PLHIV prioritize livelihood acquisition over health negatively impacting adherence to treatment and retention in care. |
| 56 | Treibich et al., 2022  (Malawi) | Cross-sectional | Drought | To analyze the effects of drought on risky sexual behavior and HIV prevalence rates | HIV  (unmarried women 15-49 years) | Transactional sex plays a role in drought-HIV relationship. Drought increases HIV prevalence by 15%. |
| 57 | Orievulu and Iwuji, 2022  (South Africa) | Qualitative using key informant (KII) and telephone interviews | Drought | To assess the institutional responses to drought impacts on HIV. | HIV  (professionals in health and disaster management sectors) | Coordination of government institutions for service delivery and interventions are weakened during drought. This is worsened by complex reporting systems and poor interdepartmental collaboration. |
| 58 | Anthonj et al., 2015  (Namibia) | Qualitative study using FGDs and KIIs & IDIs | Floods | To assess the impact of flooding on PLHIV and HIV service provision. | HIV  (PLHIV) | Flooding cuts access to facilities, increases cost of transport, disrupts access to ARV treatment, and home-based care services.  Flooding reduces HIV testing, PMTCT and ART care due to social disruption and fear of stigma. Hunger, social disruption and displacement increase during floods resulting in increases in risky sexual behavior and vulnerabilities to violence. |
| 59 | Mroz et al., 2023  (Zambia) | Cross-sectional | Floods | To assess impact of foods on women’s access to maternal health services. | MNCH  (women of reproductive age) | Floods impact women’s access to maternal health services by affecting walking duration, motorized movements and isolate women from reaching health facilities. |
| 60 | Fahey et al., 2019  (South Africa) | Cohort study | Rainfall seasonality | To assess seasonal variation in nutrition and healthcare access of pregnant women and infants. | MNCH  (pregnant women 18+ years) | ANC attendance is lowest (≤ 4), birth weight decreases during the rainy seasons. |
| 61 | Stone et al., 2020  (Mozambique) | Quantitative and retrospective | Rainfall seasonality | To investigate the associations between the rainy season and the utilization of maternal health services. | MNCH  (pregnant women) | There are lower counts of institutional deliveries during rainy season.  The effects of seasonal rainfall are significant for deliveries than for ANC visits. |
| 62 | Trickey et al., 2023  (South Africa, Zambia, Zimbabwe, Lesotho, Malawi, Mozambique) | Quantitative and retrospective | Rainfall changes | To assess the association between decreased rainfall and adverse outcomes among PLHIV on antiretroviral treatment (ART). | HIV  (PLHIV on ART) | Higher mortality occurs among PLHIV and living in areas with depressed rainfall.  Higher odds of unsuppressed viral load among people on ART and living areas with depressed rainfall. Fewer PLHIV visited HIV centers located in areas with below normal rainfall. |
| 63 | Andriano 2023  (selected SSA countries) | Cross-sectional | Heat waves | To assess the effects of exposure to heatwaves on child birthweight. | MNCH  (children) | Heat reduces gestational age and increases probability of pre-term delivery. |
| 64 | Munala et al., 2023  (Uganda  Zimbabwe  Mozambique) | Cross-sectional study | EWE (drought and floods) | To explore the relationship between extreme weather and intimate partner violence. | IPV  (partnered and married women 15-49 years) | EWE increases vulnerability of women and girls to IPV through loss of income, and unemployment. EWE is significantly associated with reported cases of IPV in the three countries. Economic burden and displacements that result from EWE are the exposure pathways for high IPV prevalence. |
| 65 | Orievulu et al., 2022  (South Africa) | Case study | Drought | To examine the economic, social, and demographic impacts of drought on PLHIV | HIV  (a rural community with high HIV prevalence) | Drought negatively affects HIV treatment adherence through disrupted income, livelihoods and food systems, and forced movements.  Drought worsens vulnerabilities linked to poverty and unemployment with implications for PLHIV. |
